# Clients’ Satisfaction With The Services Of An Outpatient Pharmacy At A University Hospital In Nigeria: A Cross-Sectional Study

**DOI:** 10.1101/2024.11.28.24318137

**Authors:** Mercy Chisom Agu, Uzochukwu Emmanuel Chima, Oluebubechukwu Praise Eze, Stanley Ndubuisi Maduekwe, Victor Udochukwu Okafor, Ozioma Maryfrances Chibuoke, Chioma Bertha Nwankwo, Christabel Ogechukwu Okoye, Amauche Pearl Ngige, Desirée Chimdimma Chigbo, Maureen Ogochukwu Akunne

## Abstract

**Background:** Pharmaceutical care involves the pharmacist’s responsibility to optimize medication therapy and improve patient outcomes. Hence, patient/client satisfaction is regarded as an important measure of the effectiveness of pharmaceutical services in achieving treatment objectives. This study assessed the level of client satisfaction with the services of an outpatient pharmacy at a university hospital in Nigeria.

**Method:** An institution online-based cross-sectional study was conducted among clients who visited the University Medical Centre at the University of Nigeria Nsukka, Nsukka campus within the period of 15^th^ September to 15^th^ October 2023. A 2-section, 30-item validated, semi-structured, interviewer-administered questionnaire was used in the study. Descriptive statistics were used to describe all the study variables while independent sample *t*-test and one-way ANOVA were used to determine the mean difference in satisfaction level among various sociodemographic characteristics. The statistical significance level was set at *p<*0.05.

**Results:** Out of the 166 participants in the study, the majority were students (n = 111, 66.9%), and receiving free medications (n = 117, 70.5%). The overall mean satisfaction score for pharmacy services was 3.11 out of a maximum 5.00 score. Notably, mean scores for most of the items exceeded 3.00 with the highest mean score obtained for the items: *“The privacy of my conversations with the pharmacist”* (3.59), and *“The fairness of cost of medications in the pharmacy”* (3.46) while the lowest score for the items: *“The way my pharmacist works together with my doctor to make sure my medications are the best for me”* (2.50) and *“The availability of medications that are prescribed to me in the pharmacy”* (2.63). First-time visitors reported significantly higher levels of satisfaction than return visitors (3.77 (0.686) vs 3.07 (0.637), *p*=<0.001)].

**Conclusion:** The study revealed a moderate level of satisfaction among the clients of the outpatient pharmacy represented in this study. The reasons for this level of satisfaction lay a foundation for the improvement of pharmacy services.

## Introduction

Good Pharmacy Practice (GPP) serves as the cornerstone for ensuring the delivery of high-quality pharmaceutical care services. Defined by established standards and guidelines, GPP sets the stage for optimal patient outcomes and satisfaction [1]. According to the World Health Organization guidelines, it required that a pharmacist’s priority is the welfare of patients; helping patients make the best use of medicines as a main activity; contributing to rational and economic prescribing and dispensing; and pharmacy services objectives should be relevant and properly communicated to patients [2].

Pharmaceutical care involves the pharmacist’s responsibility to optimize medication therapy and improve patient outcomes. To ensure the implementation of GPP with the current philosophy of pharmaceutical care, its quality should be assessed [2]. As a performance measure, patient satisfaction has been defined as the personal evaluation of health care services and providers [3]. It is a multidimensional construct that reflects the type and quality of service provided by healthcare providers, how well it is delivered, and the extent to which the expectations and needs of patients are met [3]. Satisfaction pertains to the contentment experienced by clients in response to the services provided, while the term outpatient signifies healthcare services delivered to individuals without requiring an overnight stay.

Therefore, patient satisfaction is an essential component of healthcare service quality and pharmaceutical care service. It encourages patients to improve their compliance with medications and to seek pharmaceutical care services from the same healthcare provider [4]. Adherence to GPP principles enhances the delivery of patient-centric care. This integration is marked by a focus on patient needs, safety, and effective communication, thereby fostering positive outcomes and leading to patient satisfaction [5].

Various tools have been employed to quantify the degree of patient satisfaction with pharmacy services [3]. Globally, numerous studies on patients’ satisfaction with pharmacy services provided to them have been carried out. Among these, a study conducted in Palestine highlighted the potential for enhancing patient satisfaction during the evening shift. The study suggested the implementation of a robust system and integrating technological advancements such as electronic prescribing and medication management systems for continuous evaluation and improvement of pharmaceutical care services within hospitals to ensure optimal patient care quality [4]. Another study in Saudi Arabia reported on the levels of satisfaction of clients with the services of hospital pharmacy [6]. Studies conducted in public hospitals in Eastern Ethiopia revealed notably low levels of patient satisfaction with pharmacy services [7]. Also, a study conducted at Woldia Comprehensive Specialized Hospital (WCSH), Ethiopia, demonstrated that only about half of the patients reported satisfaction with the pharmacy service [8].

In Nigeria, a study at the University of Benin Teaching Hospital unveiled subpar levels of patient satisfaction with current pharmaceutical services [9]. Also, a study conducted across eight public secondary and tertiary hospitals in Northwestern Nigeria suggested relatively high levels of respondent satisfaction with medication counseling, despite limited understanding of their prescribed medications [10]. As shown by scarce evidence of literature on the self-reported opinions of clients towards the pharmacy services received at the outpatient pharmacy at the University Medical Center located at the University of Nigeria Nsukka, Nsukka campus, it is pertinent that such study be carried out. Therefore, this study is aimed at assessing the level of the satisfaction of clients with the services of the outpatient pharmacy at the University of Nigeria Nsukka Medical Centre located at the Nsukka campus.

## Methods

### Study settings and design

This institution online-based cross-sectional study was conducted at the outpatient pharmacy unit of the University Medical Centre, University of Nigeria, Nsukka, Enugu state, Nigeria. The University Medical Centre is as old as the University and offers services at the level of Primary/Secondary health care delivery. Serious cases beyond the scope of the center are referred to the Teaching Hospital for expert management. It caters for the health needs of students, staff, and inhabitants of the host community. Students and staff are registered for a health insurance scheme, which takes care of the health needs covered by the scheme.

### Study participants and sampling

The study was conducted amongst patients who attend the medical center at the University of Nigeria Nsukka (UNN), Nsukka campus. Random sampling was adopted for this study. People who were willing and who had their prescriptions filled in at the medical center’s outpatient pharmacy, aged eighteen years or older, were allowed to participate in the study. The objective was to interview 200 clients in total over the 30-day data-collecting period. Cochran’s formula was used to determine the sample size to ensure a reliable margin of error and confidence level.

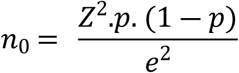

Using a Z-score of 1.96 (for a 95% confidence level) and an assumed proportion (p) of 0.5, a sample size that can accurately capture trends within the medical center’s patient population was provided. This number is also practical within the 30-day data collection period, given time and resource constraints. Every fifth client was approached to participate in the study as part of the selection procedure. This was to ensure unbiased sampling.

### Data collection instrument and process

The survey was conducted for 1 month (15^th^ September to 15^th^ October 2023). The study instrument employed in this study was a 2-section, 30-item structured self-administered questionnaire as adapted from a previous study [11]. This instrument had been used to assess the level of satisfaction of clients with the services of the pharmacies in governmental hospitals of Addis Ababa. The instrument was assessed for its reliability in the study it had been adopted from [12] and had Cronbach’s alpha of 0.9. The first section collected the socio-demographic information of the clients, and the second section contained questions assessing the level of satisfaction of clients with the services provided at the outpatient pharmacy at the University Medical Center, UNN. The second section contained twenty-one (21) questions measured on a 5-Likert scale; “1” representing “*very low*” with “2”, “3”, “4”, and “5” representing “*low*”, “*moderate*”, “*high*”, and “*very high*” respectively. The data collecting instrument was pre-tested on several customers prior to the actual data collection; these clients were later removed from the final analysis. Four lead investigators interviewed patients after they had their orders or prescriptions filled in at the pharmacy to gather data. These investigators were appropriately trained in the tool and methods in terms of approaching clients and getting their informed consent for interviews before the data collection process commenced.

### Data entry and analysis

The pre-validated questionnaire was transferred into a Google form and the link was sent to the respondents. After the data collection period, the Google form link was deactivated to stop further response. The response was downloaded as an Excel sheet and was recoded, cleaned and exported to Statistical Packages for Social Sciences (SPSS) version 27. Frequencies, percentages, means, and standard deviation were used to describe all the socio-demographic characteristics and level of satisfaction of the clients. Inferential statistics such as independent *t* test was used to assess the difference mean satisfaction between sex, payment, and patronage. Conversely, a one-way ANOVA was employed to examine the mean difference in client satisfaction levels with respect to age groups and educational status. Statistical significance was set to *p*<0.05 in all the statistical analysis.

### Ethical approval

Ethical approval was obtained from the Research Ethics Committee, University Medical Centre, UNN, Enugu State, Nigeria. The goal of the study was explained to each client, and their agreement to take part in the study was requested. They only participated in the study after a verbal consent was obtained from them. The choice of verbal consent was made because of the available short time meet with the study participants and the consent was documented in the Google form. Furthermore, the research team only used the data they had gathered for the study; patient identifiers were not used in any way. All data that was collected were handled with utmost confidentiality.

## Results

### Socio-demographic characteristics of respondents

Table I shows the socio-demographic representation of the respondents. Most of the respondents were female (n = 122, 73.5%) with a greater proportion within 18-29 age group (n = 105, 63.3%). Most of the respondents had an educational status (n = 147, 88.6%). Among the respondents, students (n = 111, 66.9%) constituted majority of the respondents followed by government employees (n = 34, 20.5%). Repeat visits constituted the highest proportion of patronage (n = 153, 92.2%) and most of the clients’ sought services for themselves (n = 140, 84.3%). In the payment status category, those getting free medication formed a higher proportion (n = 117, 70.5%) of the study participants.

**Table 1:**
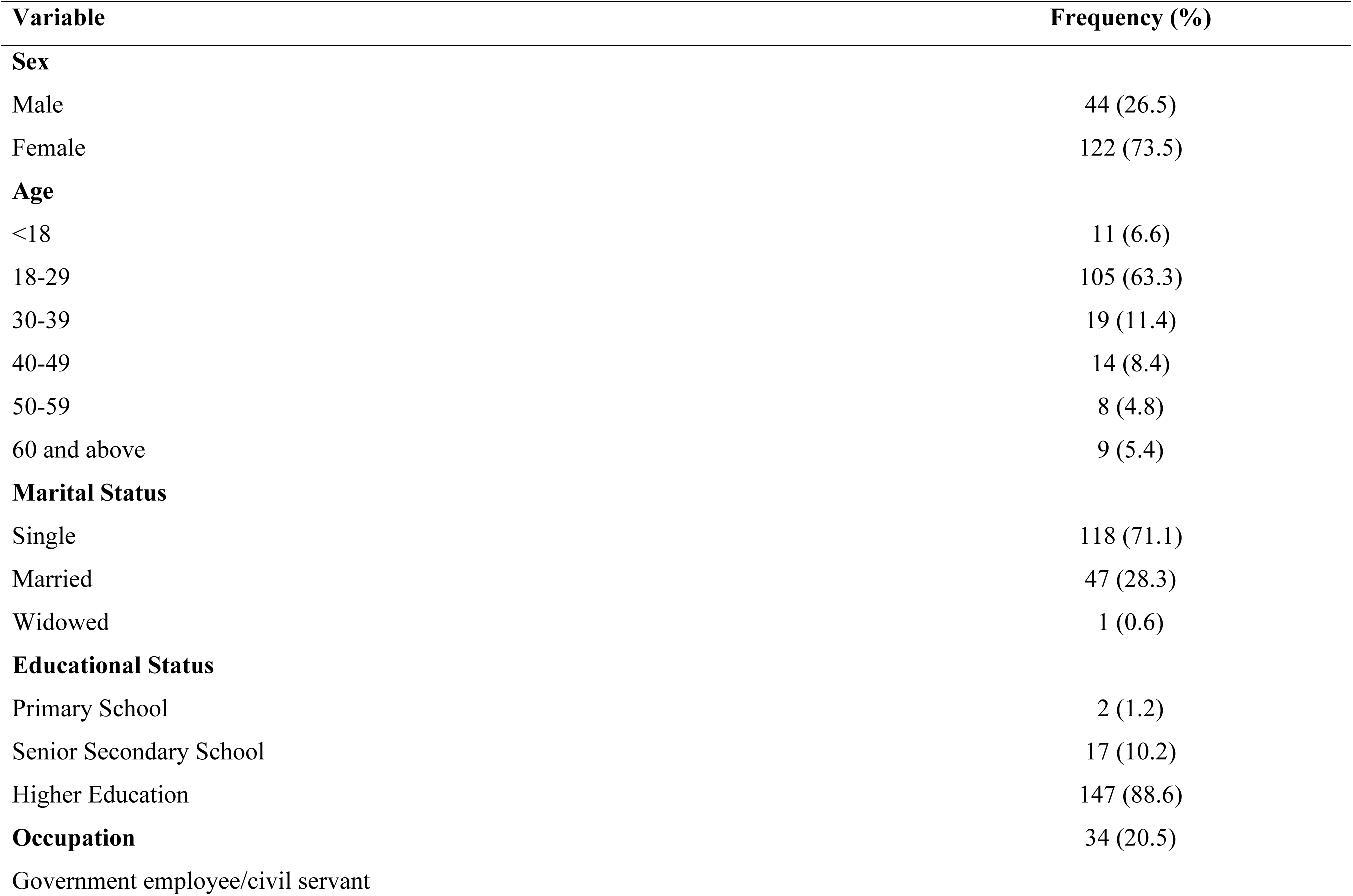
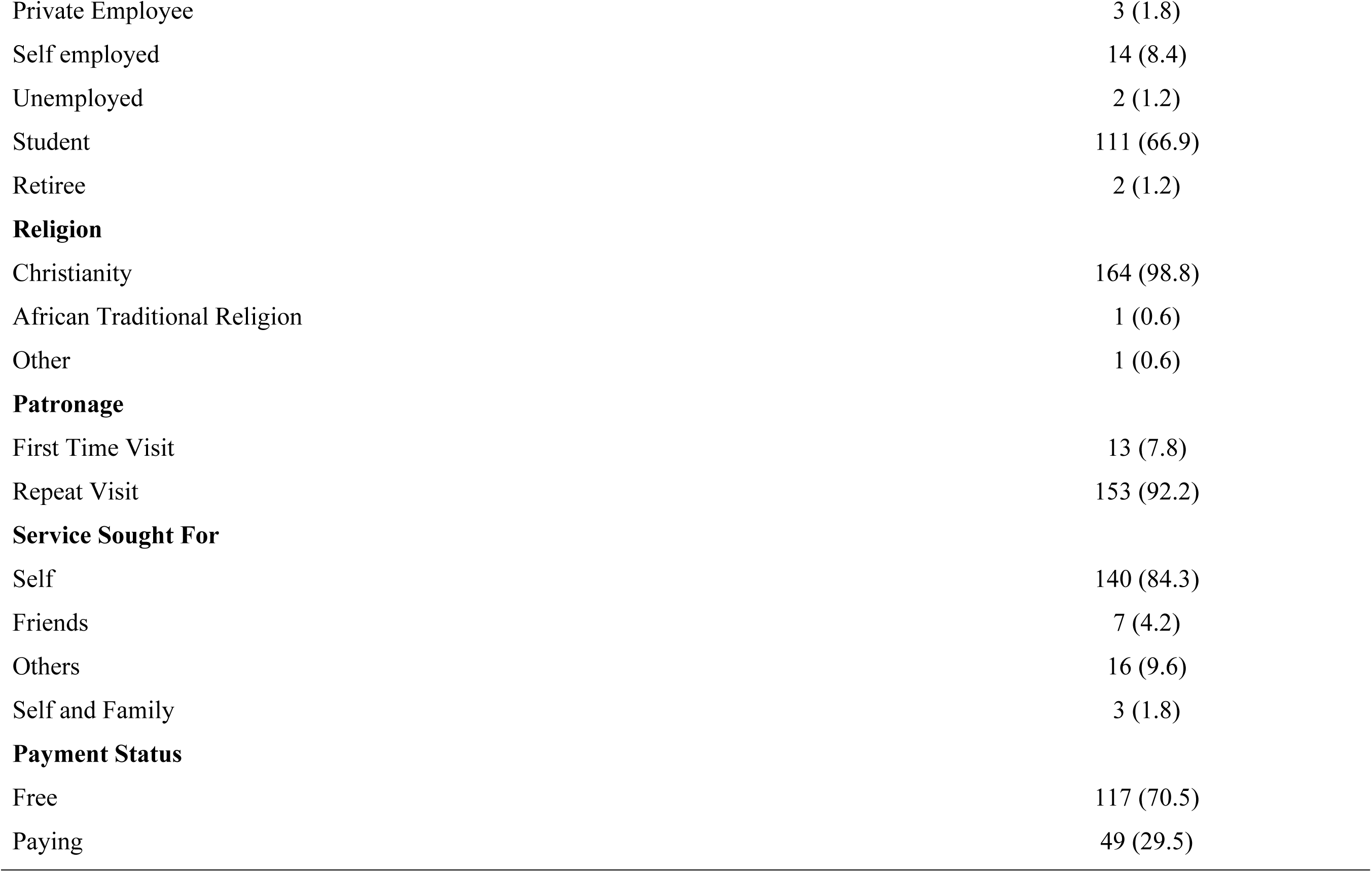
Socio-demographic Characteristics of Respondents.

### Overall satisfaction of clients with specific aspects of pharmaceutical care

Table II summarizes the satisfaction scores for each item in the study instrument. All respondents provided feedback on the 21 items they were instructed to rate. The overall mean satisfaction score given by the respondents for the pharmaceutical services rendered to them was 3.12 out of a maximum of 5.00 score. Majority of the mean scores obtained for each item on the questionnaire were above 3.00. The maximum scores were obtained for parameters: *“The privacy of my conversations with the pharmacist”* (3.59), and *“The fairness of cost of medications in the pharmacy”* (3.46) while the lowest ratings were given for parameters including: *“The way my pharmacist works together with my doctor to make sure my medications are the best for me”* (2.50) and *“The availability of medications that are prescribed to me in the pharmacy”* (2.63).

**Table 2:** Overall level of satisfaction of clients with specific aspects of pharmaceutical care.

| **Variable** | **Very Low**<br>**(%)** | **Low**<br>**(%)** | **Moderate**<br>**(%)** | **High**<br>**(%)** | **Very high**<br>**(%)** | **Mean (SD)** |
| --- | --- | --- | --- | --- | --- | --- |
| The pharmacy professional's interest in my health | 12 (7.2) | 15 (9.0) | 78 (47.0) | 49 (29.5) | 12 (7.2) | 3.20 (0.963) |
| The professionalism of all the pharmacy staff | 8 (4.8) | 27 (16.3) | 74 (44.6) | 49 (29.5) | 8 (4.8) | 3.13 (0.911) |
| The courtesy and respect shown to me by the pharmacy staff | 13 (7.8) | 16 (9.6) | 65 (39.2) | 60 (36.1) | 12 (7.2) | 3.25 (1.001) |
| The privacy of my conversations with the pharmacist | 6 (3.6) | 12 (7.2) | 52 (31.3) | 70 (42.2) | 26 (15.7) | 3.59 (0.960) |
| How well the pharmacist explains possible side effects | 30 (18.1) | 40 (24.1) | 44 (26.5) | 43 (25.9) | 9 (5.4) | 2.77 (1.180) |
| The promptness of prescription medication service | 12 (7.2) | 31 (18.7) | 66 (39.8) | 39 (23.5) | 18 (10.8) | 3.12 (1.066) |
| The care the pharmacy professional takes while supplying my medications | 14 (8.4) | 25 (15.1) | 53 (31.9) | 56 (33.7) | 18 (10.8) | 3.23 (1.101) |
| The fairness of cost of medications in the pharmacy | 10 (6.0) | 18 (10.8) | 57 (34.3) | 48 (28.9) | 33 (19.9) | 3.46 (1.110) |
| The amount of time the pharmacy professional spends with me | 15 (9.0) | 29 (17.5) | 75 (45.2) | 39 (23.5) | 8 (4.8) | 2.98 (0.984) |
| The clarity of the pharmacy professional's instructions about how to take my medication | 10 (6.0) | 23 (13.9) | 52 (31.3) | 50 (30.1) | 31 (18.7) | 3.42 (1.124) |
| The information the pharmacist gives me about the proper storage of my medication | 30 (18.1) | 40 (24.1) | 43 (25.9) | 38 (22.9) | 15 (9.0) | 2.81 (1.235) |
| How well the pharmacy professional answers my questions | 15 (9.0) | 25 (15.1) | 63 (38.0) | 48 (28.9) | 15 (9.0) | 3.14 (1.073) |
| The information the pharmacy professional gives me about the results I can expect from my medication therapy | 26 (15.7) | 40 (24.1) | 49 (29.5) | 39 (23.5) | 12 (7.2) | 2.83 (1.170) |
| The way my pharmacist works together with my doctor to make sure my medications are the best for me | 37 (22.3) | 48 (28.9) | 47 (28.3) | 29 (17.5) | 5 (3.0) | 2.50 (1.111) |
| The amount of time I spend waiting for my prescription to be filled | 13 (7.8) | 23 (13.9) | 59 (35.5) | 49 (28.5) | 22 (13.3) | 3.27 (1.102) |
| The availability of medications that are prescribed to me in the pharmacy | 34 (20.5) | 44 (26.5) | 48 (28.9) | 30 (18.1) | 10 (6.0) | 2.63 (1.173) |
| The clarity of the label on the medication supplied to me | 6 (3.6) | 27 (16.3) | 59 (35.5) | 49 (29.5) | 25 (15.1) | 3.36 (1.040) |
| My feelings of the quality of medication dispensed to me | 14 (8.4) | 25 (15.1) | 58 (34.9) | 50 (30.1) | 19 (11.4) | 3.21 (1.100) |
| The overall cleanliness and comfort of the waiting area | 8 (4.8) | 31 (18.7) | 64 (38.6) | 46 (27.7) | 17 (10.2) | 3.20 (1.016) |
| The location of the pharmacy relative to other service areas | 9 (5.4) | 25 (15.1) | 57 (34.3) | 56 (33.7) | 19 (11.4) | 3.31 (1.037) |
| My pharmacy services overall | 9 (5.4) | 22 (13.3) | 81 (48.8) | 39 (23.5) | 15 (9.0) | 3.17 (0.960) |

Among the items rated by the respondents, the majority (76.1%) were rated to be of ‘*moderate* (3.00) level of satisfaction by nearly half (48.8%) of the total respondents. In addition, in more than half (61.9%) of the items rated, only less than 22.3% of the total respondents rated the items to be of ‘*very low* (1.00)’ level of satisfaction. On the other hand, less than half (47.6%) of the items to be rated, each was rated to be a *very high* (5.00) level of satisfaction by only less than 20% of the total respondents (Table II).

### Difference in satisfaction level among respondents

In this study, the difference in the mean satisfaction level of the clients of the pharmacy involved in the study was evaluated with respect to the sociodemographic characteristics. The independent samples *t-*test was carried out on some of the sociodemographic variables including sex of respondents, patronage, service sought for and payment status. Clients who were visiting the pharmacy for the first time were shown to have a higher level of satisfaction when compared to clients who were there on return visits [3.77 (0.686) vs. 3.07 (0.637), *p*=<0.001)] (Table 3).

**Table 3:** Independent samples *t*-test of differences in the mean satisfaction level of clients by socio-demographic characteristics.

| Variable | Mean (SD) | *p*-value |
| --- | --- | --- |
| **Sex** |  |  |
| Male | 3.21 (0.545) | 0.287 |
| Female | 3.09 (0.704) |  |
| **Patronage** |  |  |
| First time visit | 3.77 (0.686) | <0.001 |
| Repeat visit | 3.07 (0.637) |  |
| **Service sought for** |  |  |
| Self | 3.09 (0.651) | 0.596 |
| Friends | 3.36 (1.283) |  |
| **Payment Status** |  |  |
| Free | 3.09 (0.692) | 0.269 |
| Paying | 3.21 (0.599) |  |

A one-way ANOVA test was carried out on different socio-demographic characteristics of clients in the study including age, marital status, educational status, occupation and religion. Although respondents aged 40-49 were more satisfied than other represented age groups [3.42 (0.527) vs.

3.07 (0.299), *p*=0.235], no statistical significance was found in this difference. Based on the results obtained, there were no statistically significant differences among other group means. (Table 4).

**Table 4:** One-way ANOVA of the differences in the mean satisfaction level of clients by socio-demographic characteristics.

| Variable | Mean (SD) | *p*-value |
| --- | --- | --- |
| **Age (in years)** |  |  |
| <18 | 3.40 (0.586) | 0.235 |
| 18-29 | 3.03 (0.735) |  |
| 30-39 | 3.16 (0.490) |  |
| 40-49 | 3.42 (0.527) |  |
| 50-59 | 3.07 (0.299) |  |
| 60 and above | 3.27 (0.519) |  |
| **Marital Status** |  |  |
| Single | 3.08 (0.696) | 0.355 |
| Married | 3.24 (0.584) |  |
| Widowed | 2.95 - |  |
| **Educational Status** |  |  |
| Primary School | 3.55 (0.505) | 0.278 |
| Senior Secondary School | 3.32 (0.517) |  |
| Higher Education | 3.09 (0.681) |  |
| **Occupation** |  |  |
| Government employer/civil servant | 3.23 (0.500) | 0.611 |
| Private Employee | 2.63 (1.296) |  |
| Self employed | 2.94 (0.650) |  |
| Unemployed | 3.26 (0.303) |  |
| Student | 3.12 (.696) |  |
| Retiree | 3.10 (1.145) |  |
| **Religion** |  |  |
| Christianity | 3.12 (0.663) | 0.137 |
| African Traditional Religion | 3.90 - |  |
| Other | 2.04 - |  |

## DISCUSSION

This study assessed the satisfaction level of clients with the services of an outpatient pharmacy at the University Medical Centre, University of Nigeria Nsukka, Nsukka campus. The respondents in this study had an overall mean satisfaction level of 3.12. The finding of this study was lower when compared to the reported mean level of a study conducted in Saudi Arabia [13] with overall mean score of 4.55. The contrast in client-reported satisfaction levels in both studies could be explained by the differences in healthcare infrastructure and patient populations. In addition, the findings of this present study reported a mean satisfaction level that is higher than the results of two studies conducted in teaching hospitals in Lagos, Nigeria [14] and Ethiopia [15] which reported slightly above average satisfaction among clients. This indicates that the practice of pharmaceutical care in this study setting may be more advanced or better structured when compared to those in Lagos, Nigeria and Ethiopia. The reported mean level of satisfaction was found to be comparable with the results of [16] where the overall mean satisfaction of respondents towards pharmacy services was 3.13. The consistency in overall mean satisfaction levels between the current study and [16] can be attributed to the fact that both outpatient pharmacies likely maintain a consistent standard of service quality or that both outpatient pharmacies provide a similar range of services hence having clients that are likely to have similar expectations and experiences regardless of the specific location. Similarly, more than half of the respondents reported dissatisfaction with the medication counselling service offered by the pharmacy professionals in Nigeria [16, 17].

The maximum satisfaction scores of respondents in this study were obtained in the items: *“privacy of conversations with the pharmacists”* and *“fairness of cost of medications in the pharmacy”*. This is different from the *“clarity of pharmacist’s instruction”* and *“pharmacist’s interest in my health”* which was rated highest in a study conducted at the University of Nigeria teaching Hospital, Ituku-Ozalla to evaluate the satisfaction level of patients with pharmaceutical care services [18]. However, it is similar to the results of the study conducted at an outpatient pharmacy in Spain [19] which reported pharmacist’s skills and confidentiality as the parameters that received highest rating. This highlights specific strengths within the outpatient pharmacy service delivery. On the other hand, the items: *“the way the pharmacist works with the doctor to make sure my medications are the best for me”* and *“availability of medications in the pharmacy”* had lowest ratings. This results, however contradicts the results of the study conducted in Ethiopia [11] which reported *“the information the pharmacist gives me about the proper storage of your medication”* and *“how well the pharmacist explains possible side effects”* as the items with lowest rating. The items rated lowest were an indication that despite the strengths of the outpatient pharmacy, there were areas for potential improvement. It suggests the need for a collaborative approach of doctors and pharmacists towards improving therapeutic outcomes and achieving overall patients’ satisfaction as suggested by [20].

Furthermore, other items such as *“how well the pharmacist explains possible side effects”* received a notably low satisfaction mean score when compared to the overall satisfaction mean score and this corresponds to the results of the studies conducted in Ethiopia [11]; Iran [21]; and Gondar University Referral Hospital, Northwestern Ethiopia [16]. The items that assessed the clients’ satisfaction level toward medication advice such as *“the information the pharmacist gives me about the proper storage of my medication”* and *“the information the pharmacy professional gives me about the results I can expect from my medication therapy”* received low ratings. This result is similar to the result of the study conducted in Makkah, Saudi Arabia [22] and that of Semegn and Alemkere [23] where very low figures were noted towards the medication guidance given to patients. This suggests that medication guidance is an area that needs to be addressed to improve the overall patient care process. Some other studies by Sanii [24] and Abebe [16] emphasized the need for proper medication counselling by pharmacists as it has been associated with treatment satisfaction.

In this study, the results of the independent samples *t-*test revealed that there was a significant association between patronage and patient’s satisfaction. Clients who patronized the outpatient pharmacy for the first time had a statistically significantly higher level of satisfaction when compared to those who were there on return visits. While this finding is similar to the findings of the study reported by Salamatullah [22], it is in contrast with the results obtained from the study conducted at the University of Nigeria Teaching Hospital, Ituku-Ozalla [18] where the patients who were there on follow-up visits exhibited a higher satisfaction level than those who were there for the first time. The lower satisfaction level recorded by returning clients may be attributed to a persistent issue of medication unavailability in the pharmacy, an item that consistently received a low rating. This has also been reported as a source of dissatisfaction in similar studies carried out in Ethiopia [25] and Tikur Anbessa, also in Ethiopia [23] and another conducted in a tertiary health facility in Lagos, Nigeria [14] where respondents identified unavailability of medicines as a major concern in public hospitals. This indicates that this is an area that needs to be considered as the availability of medicines give credibility to the health services.

The one-way ANOVA conducted on sociodemographic characteristics in this study revealed no statistically significant differences among various socio-demographic characteristics of the clients hence, sociodemographic factors of respondents did not affect the level of satisfaction. This lack of statistical significance may be attributed to the relatively homogeneous composition of the sample population in terms of age, marital status, education, occupation, and religion. This means that the study might have predominantly included participants with similar age ranges and other sociodemographic characteristics which could result in limited variability within sociodemographic detail.

### Strength And Limitations

To our knowledge, this study is the first to evaluate patients’ satisfaction with pharmaceutical services at the Medical Center at University of Nigeria, Nsukka. Research on patient satisfaction with pharmaceutical care in Nigerian tertiary institutions is scarce conferring a uniqueness to this study. But our study was not without any limitation. First, the study may be prone to sampling bias even as students form many of the end users of the pharmacy services rendered, other demographics were under-represented. Also, since the study was conducted in one setting, the results may not be used to make general conclusions for other pharmacies of tertiary institutions. Finally, the cross-sectional design of the questionnaire did not include a section for comments to be made by the respondents indicating that causality could not be implied.

### Conclusion

The study revealed that the clients at the outpatient pharmacy at the University Medical Center, UNN, Nigeria, expressed a moderate level of satisfaction for the pharmaceutical services rendered to them. Significant differences in mean satisfaction score were found between the payment status groups. This pattern of satisfaction level could serve as a pointer to better solutions geared towards improving the pharmacy services offered in the hospital.

## Data Availability

All relevant data are within the manuscript and its Supporting Information files.

